# A ketogenic supplement improves white matter energy supply and processing speed in mild cognitive impairment

**DOI:** 10.1101/2021.03.18.21253884

**Authors:** Maggie Roy, Mélanie Fortier, François Rheault, Manon Edde, Etienne Croteau, Christian-Alexandre Castellano, Francis Langlois, Valérie St-Pierre, Bernard Cuenoud, Christian Bocti, Tamas Fulop, Maxime Descoteaux, Stephen C. Cunnane

## Abstract

**INTRODUCTION:** White matter (WM) energy supply is crucial for axonal function and myelin maintenance. Providing ketones, the brain’s alternative fuel to glucose, is a therapeutic strategy to bypass the brain’s glucose-specific energy deficit in mild cognitive impairment (MCI). How an additional supply of ketones affects glucose or ketone uptake in specific WM fascicles in MCI has not previously been described.

**METHODS:** This 6-month interventional study included MCI participants randomized to a placebo (*n* = 16) or ketogenic medium chain triglyceride (kMCT; *n* = 17) drink. A neurocognitive battery and brain imaging were performed pre- and post-intervention. WM fascicle uptake of ketone and glucose and structural properties were assessed using positron emission tomography and diffusion imaging, respectively.

**RESULTS:** Ketone uptake was increased by 2.5 to 3.2-fold in nine fascicles of interest (*P* < 0.001) only in the kMCT group, an effect seen both in deep WM and in fascicle cortical endpoints. Improvement in processing speed was associated with WM ketone uptake globally and in individual fascicles, most importantly the fornix (*r* = +0.61; *P* = 0.014).

**DISCUSSION:** A 6-month ketogenic supplementation in MCI improved WM energy supply globally. The significant positive association with processing speed suggests that ketones may have a role in myelin integrity in MCI.

## 1. Background

White matter (WM) undergoes axonal degeneration and demyelination in Alzheimer’s disease (AD) [1, 2]. WM microstructural deterioration is also observed in specific brain regions in mild cognitive impairment (MCI) [3, 4]. AD is characterized by a chronic brain glucose deficit, which is already present in older people years before the onset of cognitive decline [5, 6]. WM energy supply is crucial for adequate axonal function, where oligodendrocytes need a considerable amount of energy for dynamic remodeling of myelin throughout life [7]. Hence, the gradual brain glucose deficit that begins during aging may lead to declining myelin energy metabolism, myelin loss, impaired network connectivity, thereby contributing to cognitive dysfunction [8].

Ketones (acetoacetate and β-hydroxybutyrate) are the main alternative energy substrate to glucose for the brain. In infants, ketones are not only a brain energy substrate but are also the primary substrates for myelin synthesis [9]. Recent results show that WM metabolic deterioration in AD and MCI is specific to glucose and does not involve ketones [10]. Rescuing the brain energy deficit by providing ketones is an emerging therapeutic strategy for aging-associated cognitive decline [11] and has shown promise in several randomized trials [12, 13]. The Brain ENErgy Fitness, Imaging and Cognition (BENEFIC) trial conducted in MCI showed improved measures of episodic memory, language, executive function, and processing speed after the 6-month ketogenic intervention [14] as well as improved cortical energy metabolism. WM analysis and its association to cognitive data from the BENEFIC trial has not previously been reported and are the focus of the present study.

Specific WM fascicles were targeted instead of the whole WM, since WM glucose hypometabolism was reported to be specific to limbic regions in AD [10]. The primary objectives of this report are to describe the impact of this same ketogenic supplement consumed for six months on ketone and glucose metabolism by WM fascicles and whether they were associated with the improved cognitive scores during the BENEFIC trial. The secondary objective is to report the impact of the ketogenic intervention on WM structural properties. WM energy metabolism was measured by positron emission tomography (PET) using ^11^C-acetoacetate (for ketones) and ^18^F-fluorodeoxyglucose (for glucose).

## 2. Methods

### 2.1 Participants

The BENEFIC trial (identification number NCT02551419 at ClinicalTrials.gov) was approved by our institutional ethical committee (CIUSSS de l’Estrie–CHUS, Sherbrooke, Quebec, Canada) and informed written consent was obtained from all participants before enrollment (Table 1). Inclusion criteria were male or female aged ≥ 55 years and the presence of objective cognitive decline (MCI), based on the criteria of Petersen *et al*. [15]. Specifically, MCI criteria were: subjective memory complaint plus cognitive impairment in one or more domains of a neuropsychological tests battery compared with appropriate normative data (≥ 1.5 standard deviation less than the mean), a Montreal Cognitive Assessment (MoCA) score of 18–26/30 or Mini-Mental State Examination (MMSE) score of ≥ 24/30, no evidence of probable or possible AD or depression and full autonomy for daily living activities [16]. Exclusion criteria are fully described in Fortier *et al*. [14]. The present cohort included both amnestic and non-amnestic MCI. Cognitive and depression scores, physical autonomy and blood assessments were within the normal range at baseline as detailed in Fortier *et al*. [14].

**Table 1.** Clinical data of participants at enrollment.

|  | **Placebo (*n* = 16)** | **kMCT (*n* = 16-17)\*** | **Intergroup**<br>***P* value†** |
| --- | --- | --- | --- |
| Age (y) | 75.4 ± 6.6 | 74.2 ± 6.3 | 0.600 |
| Gender (M/F) | 6/10 | 9/8 | 0.491 |
| ApoE4(+) (%) | 7/16 (44%) | 5/16 (31%) | 0.716 |
| Education (y) | 12.4 ± 3.5 | 13.1 ± 3.6 | 0.609 |
| MMSE (/30) | 26.7 ± 2.4 | 27.7 ± 2.3 | 0.126 |
| MoCA (/30) | 22.1 ± 2.4 | 23.1 ± 3.4 | 0.177 |

Abbreviations: kMCT, ketogenic medium chain triglyceride; ApoE4, apolipoprotein E4;
MMSE, Mini-Mental State Examination; MoCA, Montreal Cognitive Assessment score.
\* ApoE genotype unavailable for one participant.
† Statistical analysis was made using Mann-Whitney test or Fisher's exact test (Gender
and ApoE4 status).

### 2.2 Experimental design

Participants were randomized to the ketogenic medium chain triglyceride (kMCT) or placebo group [14]. Before starting the intervention, they underwent a dual-tracer PET and separate MRI acquisition of the brain, plus a neurocognitive battery. A second and final dual-tracer PET and MRI brain acquisition, and the neurocognitive battery were performed during the final week of the sixth month of the intervention (post-intervention).

The kMCT and placebo drinks were prepared as previously described [14]. Briefly, the kMCT drink was an emulsion containing 60% caprylic acid and 40% capric acid in lactose-free skim milk and provided 30 g/day of kMCT in a 250 mL bottle. The placebo drink contained high-oleic acid sunflower oil and was indistinguishable from the kMCT drink. During the first two weeks of the intervention, the volume to be consumed was increased from 50 mL/day to the final dose of 250 mL/day to facilitate adaptation.

### 2.3 Cognitive assessment

Testing and scoring cognition was double-blinded. Processing speed was assessed by the Trail Making (visual scanning, number sequencing, letter sequencing and motor speed) and Stroop Color and Word Interference tests from the Delis-Kaplan Executive Function System [17] and Digit Symbol Substitution tests from the Wechsler Adult Intelligence Scale [18]. All test scores were normalized and averaged to generate the attention and processing speed composite Z-score [14]. Tests for episodic memory, executive function and language domains are reported in Fortier *et al*. [14]. No learning effect was expected after the 6-month period for most of the selected tests; alternative versions for the two memory tests were used for the post-intervention assessment.

### 2.4 Image acquisition

The MRI protocol [10] was acquired on a 3 Tesla Philips Ingenia system (Best, the Netherlands) and included a 3D T1-weighted sequence (duration = 6 min, repetition time = 7.9 ms, echo time = 3.5 ms, matrix size = 240 × 240 × 160 mm, flip angle = 8° and 1 mm isotropic voxels), followed by high angular resolution diffusion imaging acquisition with parallel imaging SENSE 2 (duration = 11 min, repetition time = 11 s, echo time = 100 ms, matrix size = 128 × 128 × 78 mm, 1.8 mm isotropic voxels, 60 directions, *b* = 1500 s/mm^2^), and, one blip-up and one blip-down *b* = 0 s/mm^2^ acquisition to correct for distortions.

The MRI protocol was followed on the same day by a dual-tracer PET session [10, 14] on a PET/CT Philips Gemini system (Eindhoven, the Netherlands), in dynamic list mode. First, ^11^C-acetoacetate (AcAc) was injected for an acquisition of 10 min. One hour later, ^18^F-flurodeoxyglucose (FDG) was injected and the acquisition started 30 min post-injection and lasted for 30 min. A series of blood samples were taken during acquisitions and used to correct image-derived input functions [19]. Plasma glucose was measured with a glucose assay (Siemens Healthcare Diagnostics; Deerfield, IL) and plasma ketones were measured as previously described [20].

### 2.5 Image analysis

#### 2.5.1 Diffusion MRI

All image processing steps were blinded. Preprocessing and processing of diffusion-weighted images were performed as previously described [10] using the TractoFlow pipeline [21]. Diffusion tensor imaging (DTI) measures (fractional anisotropy, mean, radial and axial diffusivities) were computed and free-water-corrected [22] to reduce partial volume effects of the cerebrospinal fluid (CSF) associated with aging [23, 24]. Fiber orientation distribution functions were computed [25, 26] and the apparent fiber density, hereafter ‘fiber density’, at each fixel (a fiber element) was computed. The signal in a specific fiber direction (fixel) is proportional to the volume of axons aligned in that direction [27].

WM masks were corrected for white matter hyperintensities (WMH) using an average T1 template registration strategy [10].Local probabilistic tractography was performed to reconstruct whole-brain tractograms, robust to crossing fibers. Nine fascicles of interest, the ones generally reported with microstructural alteration in AD [3, 10], were extracted. The WM query language method [28] was used to automatically extract the two posterior segments of the cingulum (parahippocampal and posterior cingulate). The RecoBundlesX algorithm [29, 30] was used to extract the genu and splenium of the corpus callosum, arcuate, inferior fronto-occipital, inferior longitudinal and uncinate fasciculi. Due to its high curvature and location next to lateral ventricles contaminating voxels with CSF, the fornix was reconstructed using a separate automated fascicle-specific approach [10].

#### 2.5.2. PET

PET images were analyzed as previously described [10, 31] using PMOD 3.807 and its kinetic modeling tool PXMOD. Briefly, the Patlak method [32] was used to calculate the cerebral metabolic rate (µmol/100 g/min; referred to hereafter as ‘uptake’), and voxel-wise maps were generated for FDG and AcAc. Input functions and voxel-wise uptake maps were partial volume-corrected. Then, uptake maps were co-registered to diffusion MRI data as described previously [10] using Advanced Normalization Tools (ANTs).

#### 2.5.3 Tractometry

The quantification of PET (Fig. 1 and Video 1) and diffusion measures into fascicles was done using a tractometry pipeline [33]. FDG and AcAc uptake, free-water, fiber density and free-water-corrected DTI maps were inputs for the pipeline. Mean values were calculated for all fascicles of interest. Fascicles were divided into five sections to produce a fascicle profile that better captured ketone and glucose uptake in the extremities compared to the middle of the fascicles [10, 34].

**Figure 1.**
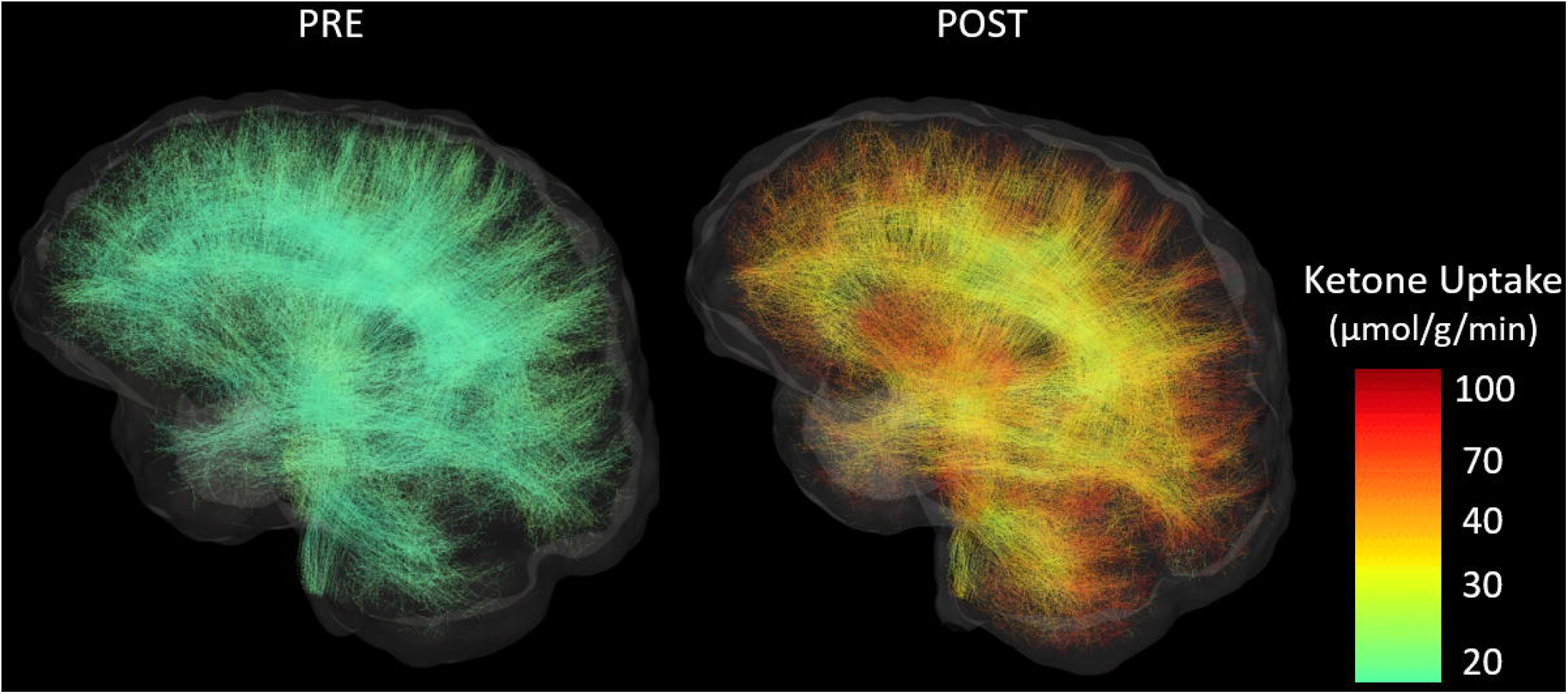
Whole-brain white matter tractograms from a sample participant in the ketogenic medium chain triglyceride (kMCT) group before (PRE) and after (POST) the 6-month intervention. Streamlines are colored according to their acetoacetate metabolic rate (µmol/g/min). Subsets of 10 000 streamlines per whole-brain tractogram are shown.

### 2.6 Statistical analysis

Complete PET and diffusion MRIs were available for *n* = 16 placebo and *n* = 17 kMCT participants. Results are presented as the mean ± standard deviation, unless otherwise mentioned. Changes in imaging measures after the 6-month intervention are expressed as percentage change from baseline. All statistical analyses were performed using SPSS 25.0 software. Since assumptions of homogeneity and normality of the variance were not met for some fascicles, non-parametric tests were used without correction for multiple comparisons. Intragroup changes (pre-vs post-intervention) were analyzed with the Wilcoxon matched-pairs signed rank test and the Mann-Whitney test was used for intergroup comparison of changes in measures (placebo vs kMCT). For fascicle profiles, pre- and post-ketone uptake measures were set as repeated measures with a general linear model to assess the effect of the intervention along the sections of each fascicle. Correlations were performed to assess a possible association between fascicle ketone uptake and the four cognitive composite scores (episodic memory, language, executive function, and attention and processing speed), as well as between both PET tracer uptake and fiber density. *P* < 0.05 was considered statistically significant in the present study.

## 3. Results

### 3.1 Clinical data

At baseline, both groups were of similar age, education, male to female ratio, ApoE4 status and education. MMSE and MoCA scores were not significantly different between groups (Table 1). All cognitive scores are presented in Fortier *et al*. [14]. Compliance was previously reported [14]. Briefly, 75% of participants completed the intervention. All completers were protocol-compliant, i.e. consumed a mean of 90% of the planned daily dose as measured by return bottle count. No severe adverse effects were observed. Some gastrointestinal effects were noted for some participants but were mostly transitory as previously reported [14].

### 3.2 Fascicle-based ketone uptake

At baseline, plasma ketones and fascicle ketone uptake were similar between groups (Table 2). After the 6-month intervention, mean ketone uptake was increased by 2.5 to 3.2-fold in all nine fascicles of interest in the kMCT group (Fig. 2), with the highest mean increase in the fornix. Mean fascicle ketone uptake was unchanged in the placebo group, and the pre- to post-intervention increase in ketone uptake was significantly higher in all fascicles in the kMCT group compared to placebo (*P* = 0.002 to 0.013; Table 2).

**Table 2.**
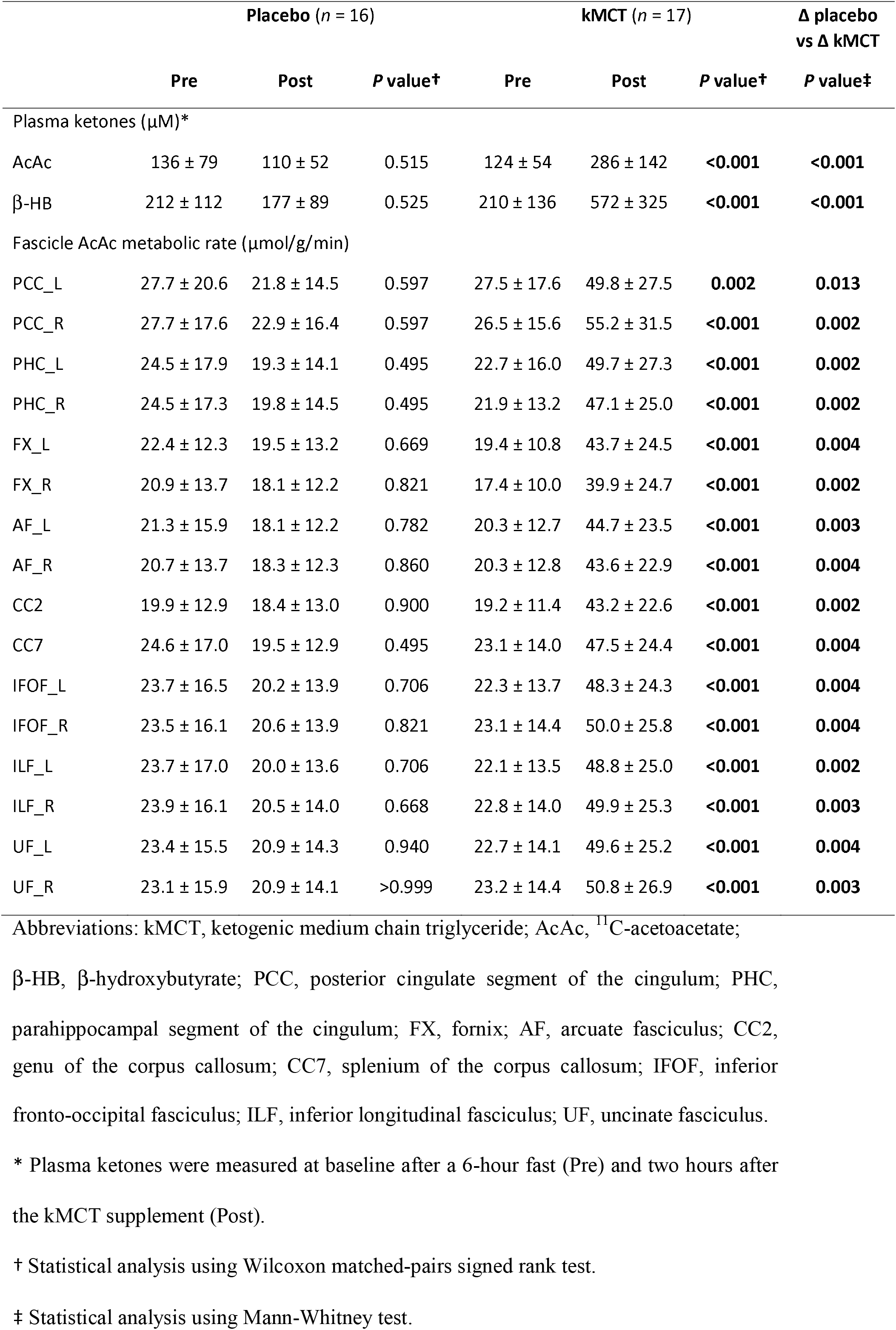
White matter fascicle-based ketone uptake (AcAc) before (Pre) and after (Post) the 6-month intervention.

**Figure 2.**
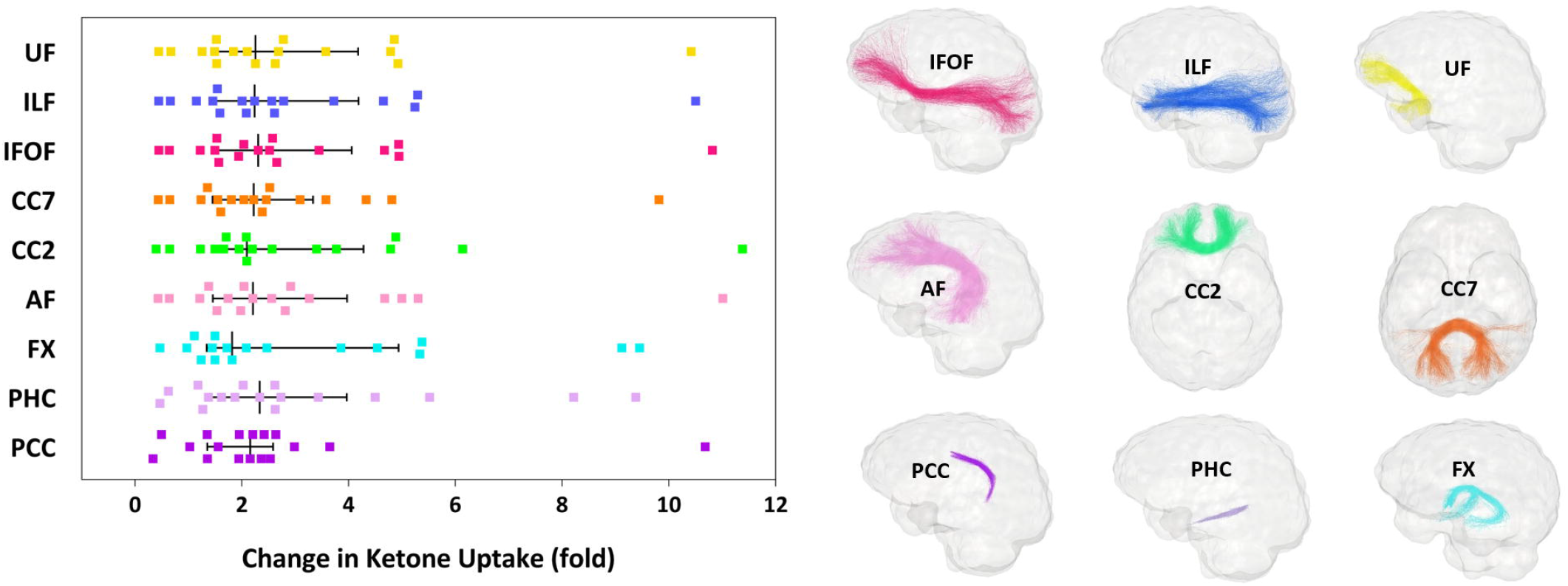
*Left*: White matter fascicle-based changes (left hemisphere only) in ketone uptake in the kMCT group after the 6-month intervention. Median ketone uptake was increased in all nine fascicles of interest after the intervention (refer to Table 2 for *P* values). Note that the median and interquartile range are illustrated for each fascicle, whereas mean values are reported in the text and tables. *Right*: Glass brain representations show the nine fascicles of interest. Fascicle colours correspond to the points in the left panel. Abbreviations: kMCT, ketogenic medium chain triglyceride; PCC, posterior cingulate segment of the cingulum; PHC, parahippocampal segment of the cingulum; FX, fornix; AF, arcuate fasciculus; CC2, genu of the corpus callosum; CC7, splenium of the corpus callosum; IFOF, inferior fronto-occipital fasciculus; ILF, inferior longitudinal fasciculus; UF, uncinate fasciculus.

Ketone uptake along fascicle section profiles was significantly increased post-intervention in the kMCT group (*P* < 0.001; Fig. A.1). The increase in ketone uptake was similar in the middle section (section 3) compared to tail section (section 5) for all nine fascicles of interest (*P* = 0.610 to 0.946). The increase of ketone uptake in the kMCT group was similar in the total WM and total cortex, 2.9 vs 2.5-fold, respectively (Table A.1; *P* = 0.865). The change in plasma ketone was positively associated with the change in total WM ketone uptake (Fig. A.2; *r* = +0.84; *P* < 0.001).

### 3.3 Association of fascicle ketone uptake with processing speed

At baseline, processing speed Z-score was similar between groups (*P* = 0.544). In the kMCT group after the 6-month intervention, the composite Z-score of processing speed was improved in direct relation to the increase in ketone uptake both in total WM (*r* = +0.52; *P* = 0.041) and in all fascicles individually (*r* = +0.46 to +0.61; *P* = 0.014 to 0.072), except for the posterior cingulate segment of the cingulum (Fig. 3 and Table A.2). In the placebo group, post-intervention changes in processing speed were inversely related to ketone uptake in total WM (*r* = −0.53; *P* = 0.043) and in individual fascicles (Table A.2), a relationship not found at baseline. Slopes between both groups were significantly different for all nine fascicles of interest (*P* = 0.003 to 0.028). The strongest association between the improvement in processing speed Z-score and the increase in ketone uptake in the kMCT group was in the fornix (*r* = +0.61; *P* = 0.014). Pre-intervention, ketone uptake in the fornix also had the strongest association with processing speed Z-score when both treatment groups were combined (*r* = +0.44; *P* = 0.010; data not shown).

**Figure 3.**
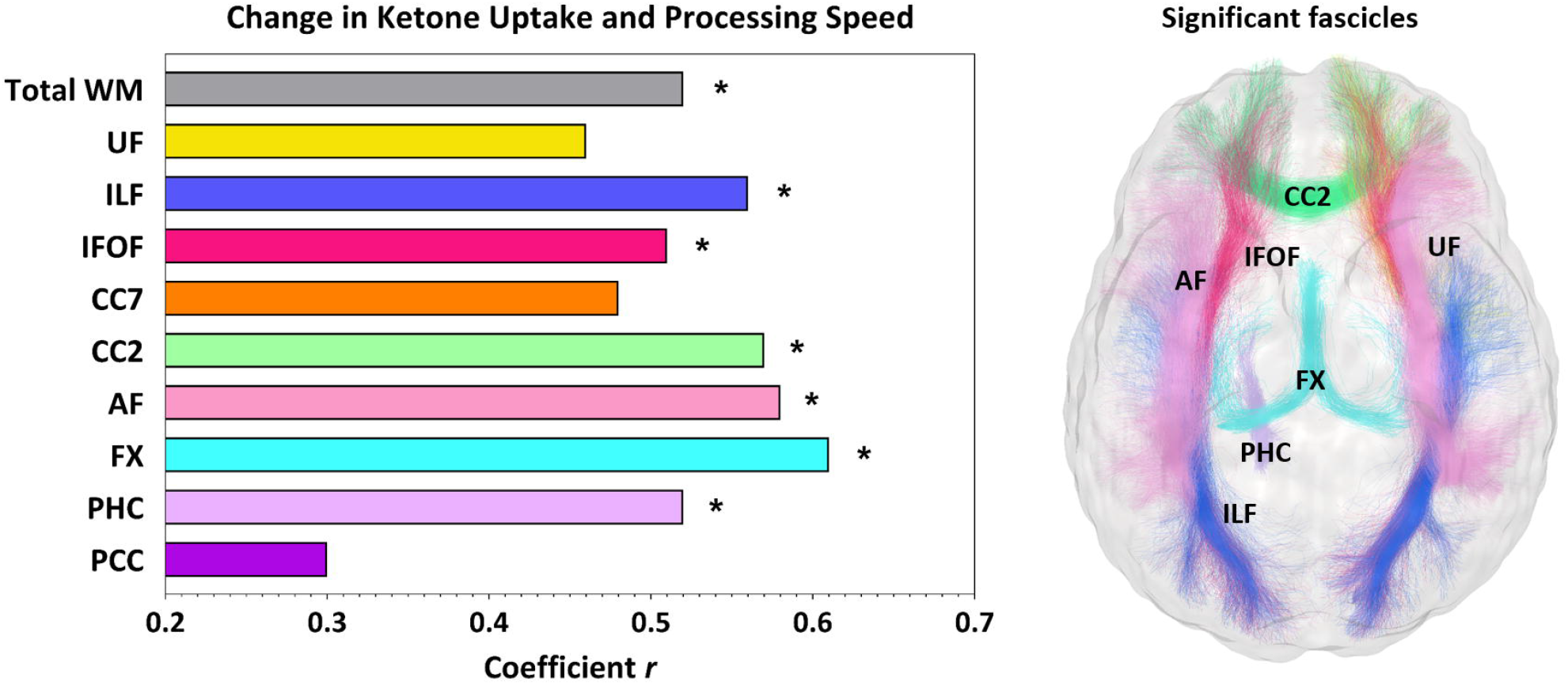
*Left*: Composite Z-score of processing speed was improved in association with the mean increase of ketone uptake in total white matter (*r* = +0.52; *P* = 0.041) and in several fascicles in the kMCT group after the 6-month intervention. Refer to Table A.2 for *P* values. * = *P* < 0.05. Fascicles from the left hemisphere are presented. *Right:* The glass brain representation shows all fascicles for which the association between change in ketone uptake and processing speed was significant. Fascicle colours correspond to the bars in the panel on the left. Abbreviations: PCC, posterior cingulate segment of the cingulum; PHC, parahippocampal segment of the cingulum; FX, fornix; AF, arcuate fasciculus; CC2, genu of the corpus callosum; CC7, splenium of the corpus callosum; IFOF, inferior fronto-occipital fasciculus; ILF, inferior longitudinal fasciculus; UF, uncinate fasciculus; WM, white matter.

Increased fascicle ketone uptake was not associated with episodic memory, language or executive function composite scores. Therefore, further analyses were restricted to measures of processing speed. In the kMCT group, improved score (decreased time taken) post-intervention on the motor speed task of the Trail Making Test was significantly inversely associated with increased ketone uptake in total WM (*r* = −0.50; *P* = 0.043) and in all fascicles individually (Table A.3). No such association was observed for the placebo group.

Post-intervention, processing speed was 14% slower on the visual scanning task of the Trail Making Test for the placebo group (pre: 30 s and post: 34 s; *P* = 0.040) but remained unchanged in the kMCT group (*P* = 0.922). In the kMCT group, improvement on the visual scanning task of the Trail Making Test was also associated with increased ketone uptake in total WM (*r* = −0.53; *P* = 0.036) and in the fornix, genu of the corpus callosum, arcuate, inferior fronto-occipital and inferior longitudinal fasciculi (data not shown).

### 3.4 Fascicle-based diffusion measures

No difference in diffusion measures pre- to post-intervention was found between the two treatments. After the 6-month intervention, the fiber density was 8-14% lower in most fascicles of both groups, except for the fornix, and parahippocampal and posterior cingulate segments of the cingulum (Table A.4). The splenium of the corpus callosum had the greatest decrease in fiber density. In contrast, fiber density was increased by 11-18% post-intervention in both groups in the left parahippocampal and posterior cingulate segments of the cingulum, respectively. Free-water-corrected radial diffusivity was significantly increased in several fascicles post-intervention in both groups but was reduced only in the posterior cingulate segment of the cingulum (*P* < 0.001 to 0.011; data not shown). Free-water-corrected fractional anisotropy was increased by 15-18% in the posterior cingulate segment of the cingulum in both groups (*P* < 0.001 to 0.003; data not shown). Post-intervention, both groups had an increase in free-water of 4-23% in all fascicles, especially the parahippocampal segment of the cingulum, except in the left posterior cingulate segment of the cingulum and right fornix (Table A.5).

### 3.5 Fascicle-based glucose uptake and association with fiber density

No significant difference was found for fascicle glucose uptake between groups at baseline (Table A.6). No difference in glucose uptake pre-to post-intervention was found between the two treatments. Post-intervention, mean glucose uptake was unchanged in all fascicles, except in the posterior cingulate segment of the cingulum where it was reduced by 9-11% in both groups (*P* = 0.009 to 0.044; Table A.6). When combined across both groups, the change in glucose uptake was positively associated with change in fiber density in the uncinate fasciculus, fornix and genu of the corpus callosum (*r* = +0.41 to +0.52; *P* = 0.003 to 0.028), but inversely in the posterior cingulate segment of the cingulum (*r* = −0.45; *P* = 0.012; Fig. 4). For these significant associations, slopes were significantly non-zero (*P* = 0.003 to 0.030; data not shown).

**Figure 4.**
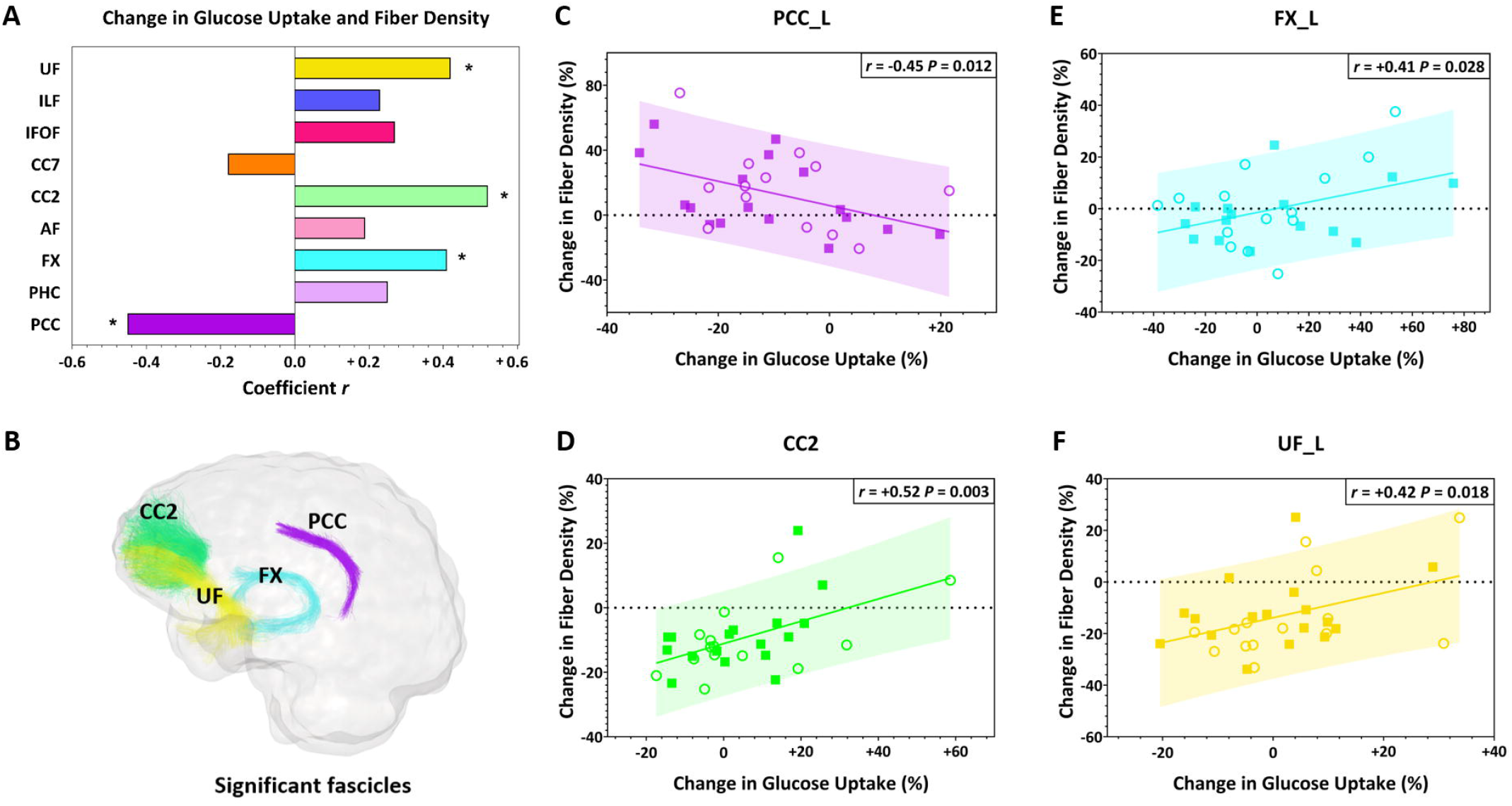
(A) After the 6-month intervention, change in mean glucose uptake was positively associated with change in fiber density in the uncinate fasciculus, fornix and genu of the corpus callosum, but inversely associated in the posterior cingulate segment of the cingulum. Data from both the placebo (○) and kMCT (□■) groups were combined because no significant treatment effect was found for these variables. * = *P* < 0.05. Fascicles from the left (L) hemisphere are presented. (B) The glass brain representation shows all fascicles for which the association was significant. Fascicle colours correspond to the bars in panel A. (C-F) Scatter plots of significant associations between change in glucose uptake and fiber density. Abbreviations: PCC, posterior cingulate segment of the cingulum; PHC, parahippocampal segment of the cingulum; FX, fornix; AF, arcuate fasciculus; CC2, genu of the corpus callosum; CC7, splenium of the corpus callosum; IFOF, inferior fronto-occipital fasciculus; ILF, inferior longitudinal fasciculus; UF, uncinate fasciculus.

## 4. Discussion

This is the first study to report the impact of a ketogenic intervention on WM structure and energy metabolism in humans. We show that a 6-month kMCT supplement improved WM energy supply, with a nearly 3-fold increase in ketone uptake, but with no significant effect on glucose uptake. The increased ketone uptake was present in all WM fascicles assessed and was as important in the deep WM as in the fascicle endpoints in gray matter. Processing speed improvement was positively associated with increased WM ketone uptake globally and in individual fascicles, most importantly in the fornix. This association suggests that ketones may have a role in myelin integrity in MCI. In MCI, increased fiber density was seen specifically in the posterior cingulum, which also had declining glucose metabolism during the 6-month study period.

In the kMCT group, ketone uptake increased over six months in all fascicles analyzed (Fig. 1, 2 and Video 1). The fornix had the highest increase in mean ketone uptake post-intervention, which may in part be attributed to its lower ketone and glucose uptake at baseline. This increase in ketone availability to the fornix may potentially be beneficial to limit the early decline of structural integrity of the fornix during aging [35]. A global 2.5-fold increase of ketone uptake was also found for the cortex (Table A.1; not significantly different from the 2.9-fold increase in total WM. Clearly, in MCI, WM avidly consumes ketones when they are available.

In the kMCT group, processing speed composite Z-score improved in association with the increase of ketone uptake in almost all fascicles and also in WM as a whole (Fig. 3). This suggests that information processing is probably linked to global structural connectivity in MCI, rather than to a specific fascicle, which agrees with previous studies in older adults [36, 37]. For specific cognitive tests, the strongest association with increased ketone uptake was for the motor speed task of the Trail Making Test, a nearly pure measure of processing speed. Improved processing speed scores may be a consequence of increased myelin density following the ketogenic intervention. Indeed, degeneration of myelin lipids associated with aging has been proposed to provide fatty acids or ketones as an energetic substrate for neurons [38]. Processing speed is directly linked to myelin integrity, as the nerve conduction of action potentials is proportional to the thickness of myelin sheaths [39]. Declining processing speed is a key deficit in multiple sclerosis [40], a disease with widespread demyelination of the WM. Processing speed may also have improved due to increased intra-axonal mitochondrial production of ATP [41], resulting from an elevated ketone supply to the brain. An important part of the energy used by the brain is to maintain information processing speed through impulse propagation [8]. In the BENEFIC trial [13], positive correlations between plasma ketones and tests of episodic memory, executive function and language were found, but not with processing speed. This reinforces the essential role of WM in processing speed, where ketones may act as an important substrate.

In contrast to the kMCT group, in the placebo group, an inverse association was found between WM ketone uptake and processing speed. Slower processing speed in the placebo group may be attributable to disease progression over the six months. Indeed, WM pathology in MCI is associated with impaired processing speed [42]. There was no significant association between ketone uptake by the fornix and memory, but there was a strong correlation with processing speed. The microstructure of the fornix has previously been reported to be associated to processing speed but not to memory in older adults [43] and in epilepsy [44], suggesting a role of the fornix beyond memory function.

The 6-month ketogenic supplementation had no effect on structural WM measures. In other studies, physical activity interventions of one year also did not impact WM microstructure in older adults [45] and MCI [46]. We show here that, as opposed to other fascicles, the fiber density of the posterior cingulum increased in both groups (Table A.4). This supports the ‘network failure’ theory [47, 48], which proposes that in the prodromal phase of AD, the posterior default mode network (a key processing hub), starts compensating for declining function of other brain networks, becomes overloaded and starts to shift its processing load to related networks, resulting in increased connectivity between the posterior default mode network and other hubs. Structural connectivity changes associated with cognitive decline may therefore follow a non-linear trajectory for some specific networks.

We show here a decline in glucose uptake over six months specifically in the posterior cingulate segment of the cingulum in both groups (Table A.6). In the kMCT group the increase of ketone uptake and decrease of glucose uptake in this fascicle were not significantly correlated (data not shown), so it seems possible that disease progression over the six months could have been linked specifically to declining glucose uptake in the posterior cingulate segment of the cingulum. The posterior cingulate segment of the cingulum had the highest glucose uptake at baseline (62% higher than the fornix), which may make this region more vulnerable to declining energy (glucose) metabolism during aging. The increase in ketone uptake post-intervention was also the lowest in the posterior cingulate segment of the cingulum. This would make it more difficult for ketones to compensate for impaired glucose uptake in posterior cingulate segment of the cingulum during MCI. Other tractography studies show that the posterior cingulum has lower glucose uptake [10] and fiber density [3] in MCI compared to controls, suggesting it is a particularly vulnerable fascicle when MCI develops.

A limitation of the present study was the small sample size for assessing the relationships to cognitive outcomes. Although WMH were eliminated for robust fascicle reconstruction, another limitation was the inability to segment these lesions and establish whether both groups had a similar cerebrovascular burden. However, in terms of cardiovascular risk factors, both groups had similar blood pressure and plasma cholesterol at baseline [14]. WMH regional load may have affected the cerebral metabolic rate of PET tracers, but the relationship between WM ketone uptake and processing speed was still highly significant. In future studies, including fluid-attenuated inversion recovery (FLAIR) images will be important for automated segmentation of WMH [49]. Because of the importance of myelin for information processing and its link to ketone metabolism, assessment of the myelin content would also be recommended in the future when ketogenic or other energetic interventions are used. The lack of specificity of radial diffusivity to myelin requires a more sophisticated MRI sequence, such as the inhomogeneous magnetization transfer data [50], more specific to myelin.

In conclusion, we show that a 6-month kMCT supplement increased ketone supply in WM fascicles in MCI, with an equivalent effect in deep WM as in fascicle gray matter endpoints. Several improved measures of processing speed were directly associated with increased WM ketone supply globally and in individual fascicles, most importantly in the fornix. Finally, these data support the idea of a posterior cingulum vulnerability at the MCI stage. The impact of a long-term ketogenic supplementation on myelin density should be investigated in order to better understand the role of ketones in myelin integrity and cognition in MCI.

## Data Availability

Original data and processed data will be made available upon request. All processing steps and
results can be reproduced using the TractoFlow
pipeline.

## Abbreviations

AD: Alzheimer’s disease
MCI: mild cognitive impairment
WM: white matter
kMCT: ketogenic medium chain triglyceride
ApoE4: apolipoprotein E4
MMSE: Mini-Mental State Examination
MoCA: Montreal Cognitive Assessment score
MRI: magnetic resonance imaging
DTI: diffusion tensor imaging
CSF: cerebrospinal fluid
WMH: white matter hyperintensities
PET: positron emission tomography
FDG: ^18^F-flurodeoxyglucose
AcAc: ^11^C-acetoacetate
β-HB: beta-hydroxybutyrate
PCC: posterior cingulate segment of the cingulum
PHC: parahippocampal segment of the cingulum
FX: fornix
AF: arcuate fasciculus
CC2: genu of the corpus callosum
CC7: splenium of the corpus callosum
IFOF: inferior fronto-occipital fasciculus
ILF: inferior longitudinal fasciculus
UF: uncinate fasciculus

## Acknowledgments

The authors wish to acknowledge the PET and MRI clinical teams at the Sherbrooke Molecular Imaging Center, Matthieu Dumont, Camille Vandenberghe, Dr. Sébastien Tremblay and Marie-Christine Morin for technical assistance, the Sherbrooke Connectivity Imaging Lab team for their help on data processing, and Dr. Alexa Pichet Binette and Dr. Sylvia Villeneuve for their assistance on data interpretation. We would also like to acknowledge the participants of the BENEFIC trial.

Financial support for the BENEFIC trial was provided by the Alzheimer Association USA (PCTR-15-328047), FRQS (FR40072), and Université de Sherbrooke. MR was funded by MITACS and Nestlé Health Science. The Université de Sherbrooke Institutional Chair in Neuroinformatics also provided funding support. Abitec provided the kMCT (Captex 355) and placebo oil. The intervention drinks for both arms were prepared under contract at INAF, Université Laval, Québec, QC, Canada.

## Notes

### Competing Interest Statement

SCC has consulted for or received travel honoraria or test products from Nestle Health Science, Bulletproof, Cerecin, and Abitec. SCC is the founder and director of the consulting company, Senotec Ltd. MD is CSO and shareholder of Imeka Solutions Inc (www.imeka.ca) and MR is consultant for Imeka Solutions Inc.

### Clinical Trial

NCT02551419

### Author Declarations

Institutional ethical committee (CIUSSS de l Estrie CHUS Sherbrooke Quebec Canada).

