## Appendices (A) for "A ketogenic supplement improves white matter energy supply and processing speed in mild cognitive impairment"

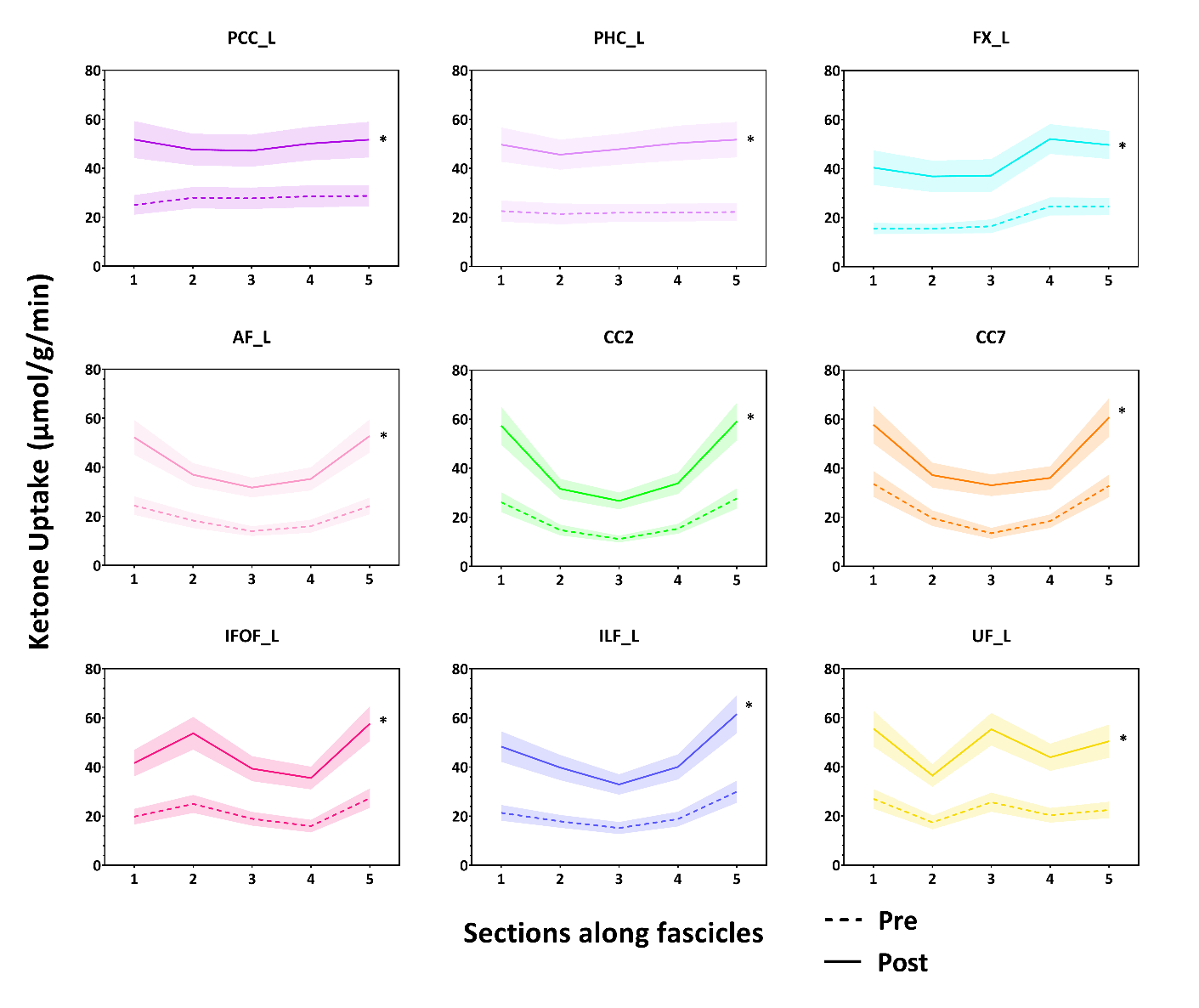
**Appendices**

**Figure A.1.** Profiles of ketone uptake along fascicle sections in the kMCT group before (Pre) and after (Post) the 6-month intervention. Ketone uptake (AcAc metabolic rate; µmol/g/min) along all five sections of all fascicles was significantly increased pre- to post-intervention (* = *P* < 0.001). Fascicles from the left (L) hemisphere are presented and colored to correspond those in Fig. 2. Abbreviations: PCC, posterior cingulate segment of the cingulum; PHC, parahippocampal segment of the cingulum; FX, fornix; AF, arcuate fasciculus; CC2, genu of the corpus callosum; CC7, splenium of the corpus callosum; IFOF, inferior fronto-occipital fasciculus; ILF, inferior longitudinal fasciculus; UF, uncinate fasciculus.

| Table A.1. Global ketone (AcAc) and glucose (FDG) uptake before (Pre) and after (Post) the 6-month intervention in the kMCT group (*n* = 17). | | | | | | | |
| --- | --- | --- | --- | --- | --- | --- | --- |
|  | **Ketone uptake** (µmol/g/min)* | | | **Glucose uptake** (µmol/100 g/min)* | | | |
|  | **Pre** | **Post** | ***P* value**† | | **Pre** | **Post** | ***P* value**† |
| Total white matter | 28.6 ± 17.5 | 61.8 ± 32.4 | **<0.001** | | 21.77 ± 3.37 | 21.75 ± 3.25 | 0.890 |
| Total cortex | 40.9 ± 22.8 | 90.5 ± 61.4 | **0.001** | | 30.94 ± 4.10 | 30.19 ± 4.01 | 0.555 |
| Whole brain | 37.1 ± 20.6 | 81.8 ± 54.4 | **0.001** | | 27.46 ± 3.65 | 26.82 ± 3.50 | 0.619 |

Abbreviations: kMCT, ketogenic medium chain triglyceride; AcAc, ^11^C-acetoacetate; FDG, ^18^F-fluorodeoxyglucose.

* Note the difference in units for ketone vs glucose uptake.

† Statistical analysis was made using Wilcoxon matched-pairs signed rank test.

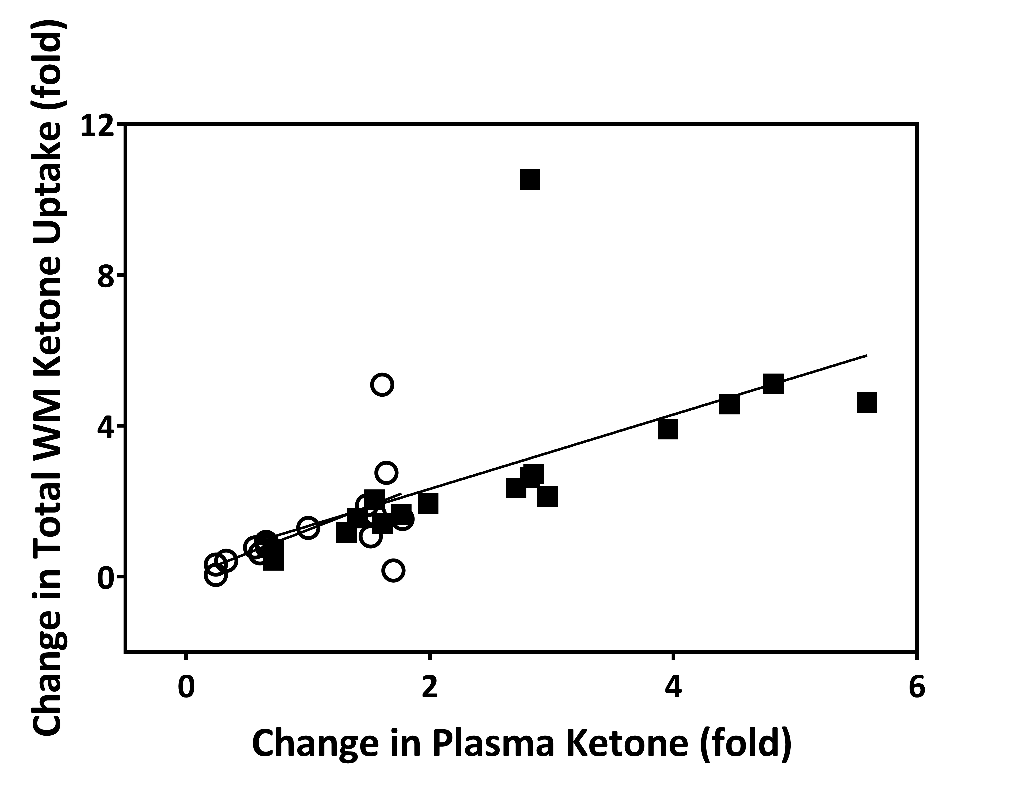

**Figure A.2.** Scatter plot of the association between change in plasma ketone (acetoacetate) and total white matter ketone uptake (*r* = +0.84; *P* < 0.001). Both groups were included in the statistical analysis: placebo (○) and kMCT (󠇥■) groups. Abbreviation: WM, white matter.

| **Table A.2. Correlation between change in ketone uptake (AcAc)* and attention and processing speed composite Z-score.** | | | | | |
| --- | --- | --- | --- | --- | --- |
|  | **Placebo** (*n* = 16) | | **kMCT** (*n* = 16)† | | **Difference between slopes**  ***P* value** |
|  | ***r* value** | ***P* value** | ***r* value** | ***P* value** |  |
| PCC_L | -0.52 | 0.051 | 0.30 | 0.263 | **0.028** |
| PCC_R | -0.55 | **0.037** | 0.48 | 0.062 | **0.024** |
| PHC_L | -0.46 | 0.083 | 0.52 | **0.041** | **0.009** |
| PHC_R | -0.51 | 0.052 | 0.48 | 0.062 | **0.004** |
| FX_L | -0.54 | **0.040** | 0.61 | **0.014** | **0.003** |
| FX_R | -0.54 | **0.038** | 0.51 | **0.047** | **0.003** |
| AF_L | -0.53 | **0.042** | 0.58 | **0.020** | **0.010** |
| AF_R | -0.53 | **0.043** | 0.57 | **0.023** | **0.009** |
| CC2 | -0.53 | **0.043** | 0.57 | **0.024** | **0.007** |
| CC7 | -0.55 | **0.037** | 0.48 | 0.063 | **0.013** |
| IFOF_L | -0.53 | **0.043** | 0.51 | **0.045** | **0.011** |
| IFOF_R | -0.51 | 0.052 | 0.53 | **0.037** | **0.011** |
| ILF_L | -0.53 | **0.042** | 0.56 | **0.026** | **0.008** |
| ILF_R | -0.53 | **0.043** | 0.52 | **0.040** | **0.009** |
| UF_L | -0.53 | **0.044** | 0.46 | 0.072 | **0.010** |
| UF_R | -0.53 | **0.042** | 0.52 | **0.042** | **0.007** |

Abbreviations: kMCT, ketogenic medium chain triglyceride; AcAc, ^11^C-acetoacetate; PCC, posterior cingulate segment of the cingulum; PHC, parahippocampal segment of the cingulum; FX, fornix; AF, arcuate fasciculus; CC2, genu of the corpus callosum; CC7, splenium of the corpus callosum; IFOF, inferior fronto-occipital fasciculus; ILF, inferior longitudinal fasciculus; UF, uncinate fasciculus.

* Fascicle AcAc metabolic rate (µmol/g/min)

† Some cognitive test results were not available for one participant.

| **Table A.3. Correlation between change in ketone uptake (AcAc)* and Trail Making Test results (motor speed task; s).** | | | | | |
| --- | --- | --- | --- | --- | --- |
|  | **Placebo** (*n* = 16) | | **kMCT** (*n* = 16)† | | **Difference between slopes**  ***P* value** |
|  | ***r* value** | ***P* value** | ***r* value** | ***P* value** |  |
| PCC_L | -0.35 | 0.188 | -0.38 | 0.134 | 0.696 |
| PCC_R | -0.31 | 0.250 | -0.52 | **0.032** | 0.529 |
| PHC_L | -0.40 | 0.133 | -0.55 | **0.023** | 0.405 |
| PHC_R | -0.39 | 0.144 | -0.53 | **0.031** | 0.354 |
| FX_L | -0.32 | 0.247 | -0.47 | 0.060 | 0.137 |
| FX_R | -0.41 | 0.129 | -0.49 | **0.046** | 0.070 |
| AF_L | -0.34 | 0.208 | -0.54 | **0.025** | 0.923 |
| AF_R | -0.35 | 0.200 | -0.53 | **0.030** | 0.913 |
| CC2 | -0.34 | 0.200 | -0.55 | **0.023** | 0.802 |
| CC7 | -0.31 | 0.250 | -0.51 | **0.037** | 0.846 |
| IFOF_L | -0.35 | 0.200 | -0.49 | **0.045** | 0.908 |
| IFOF_R | -0.37 | 0.164 | -0.52 | **0.036** | 0.963 |
| ILF_L | -0.34 | 0.207 | -0.54 | **0.025** | 0.827 |
| ILF_R | -0.35 | 0.200 | -0.53 | **0.030** | 0.800 |
| UF_L | -0.35 | 0.200 | -0.55 | **0.024** | 0.900 |
| UF_R | -0.34 | 0.208 | -0.54 | **0.028** | 0.856 |

Abbreviations: kMCT, ketogenic medium chain triglyceride; AcAc, ^11^C-acetoacetate; PCC, posterior cingulate segment of the cingulum; PHC, parahippocampal segment of the cingulum; FX, fornix; AF, arcuate fasciculus; CC2, genu of the corpus callosum; CC7, splenium of the corpus callosum; IFOF, inferior fronto-occipital fasciculus; ILF, inferior longitudinal fasciculus; UF, uncinate fasciculus.

* Fascicle AcAc metabolic rate (µmol/g/min)

† Some cognitive test results were not available for one participant.

| **Table A.4. White matter fascicle-based fiber density before (Pre) and after (Post) the 6-month intervention.** | | | | | | | |
| --- | --- | --- | --- | --- | --- | --- | --- |
|  | **Placebo** (*n* = 16) | | | **kMCT** (*n* = 17) | | | **Δ placebo vs Δ kMCT** |
|  | **Pre** | **Post** | ***P* value*** | **Pre** | **Post** | ***P* value*** | ***P* value**† |
| PCC_L | 0.40 ± 0.05 | 0.48 ± 0.12 | **0.008** | 0.39 ± 0.07 | 0.44 ± 0.14 | 0.145 | 0.510 |
| PCC_R | 0.41 ± 0.06 | 0.46 ± 0.11 | **0.015** | 0.41 ± 0.06 | 0.44 ± 0.11 | 0.120 | 0.606 |
| PHC_L | 0.27 ± 0.05 | 0.30 ± 0.08 | **0.025** | 0.29 ± 0.06 | 0.31 ± 0.06 | **0.020** | 0.487 |
| PHC_R | 0.27 ± 0.06 | 0.30 ± 0.08 | 0.175 | 0.28 ± 0.06 | 0.29 ± 0.06 | 0.890 | 0.465 |
| FX_L | 0.22 ± 0.04 | 0.22 ± 0.03 | 0.744 | 0.23 ± 0.04 | 0.23 ± 0.05 | 0.489 | 0.572 |
| FX_R | 0.21 ± 0.03 | 0.21 ± 0.03 | 0.597 | 0.22 ± 0.04 | 0.22 ± 0.04 | 0.638 | 0.599 |
| AF_L | 0.46 ± 0.04 | 0.43 ± 0.08 | 0.175 | 0.46 ± 0.06 | 0.41 ± 0.07 | **0.003** | 0.581 |
| AF_R | 0.47 ± 0.04 | 0.43 ± 0.06 | **0.033** | 0.46 ± 0.06 | 0.42 ± 0.06 | **0.005** | 0.631 |
| CC2 | 0.40 ± 0.03 | 0.36 ± 0.06 | **0.008** | 0.40 ± 0.05 | 0.36 ± 0.06 | **0.007** | 0.929 |
| CC7 | 0.63 ± 0.10 | 0.54 ± 0.08 | **0.003** | 0.61 ± 0.13 | 0.53 ± 0.08 | **0.001** | 0.736 |
| IFOF_L | 0.46 ± 0.05 | 0.43 ± 0.06 | 0.051 | 0.46 ± 0.06 | 0.42 ± 0.08 | **0.004** | 0.631 |
| IFOF_R | 0.49 ± 0.04 | 0.45 ± 0.05 | **0.008** | 0.50 ± 0.06 | 0.44 ± 0.06 | **<0.001** | 0.423 |
| ILF_L | 0.43 ± 0.05 | 0.40 ± 0.06 | **0.015** | 0.44 ± 0.05 | 0.40 ± 0.07 | **0.001** | 0.657 |
| ILF_R | 0.46 ± 0.03 | 0.42 ± 0.04 | **0.011** | 0.47 ± 0.06 | 0.42 ± 0.07 | **<0.001** | 0.292 |
| UF_L | 0.38 ± 0.04 | 0.33 ± 0.06 | **0.009** | 0.38 ± 0.04 | 0.33 ± 0.06 | **0.005** | 0.423 |
| UF_R | 0.39 ± 0.04 | 0.35 ± 0.06 | **0.009** | 0.40 ± 0.05 | 0.34 ± 0.06 | **<0.001** | 0.204 |

Abbreviations: kMCT, ketogenic medium chain triglyceride; PCC, posterior cingulate segment of the cingulum; PHC, parahippocampal segment of the cingulum; FX, fornix; AF, arcuate fasciculus; CC2, genu of the corpus callosum; CC7, splenium of the corpus callosum; IFOF, inferior fronto-occipital fasciculus; ILF, inferior longitudinal fasciculus; UF, uncinate fasciculus.

* Statistical analysis was made using Wilcoxon matched-pairs signed rank test.

† Statistical analysis was made using Mann-Whitney test.

| **Table A.5. White matter fascicle-based free-water before (pre) and after (post) the 6-month intervention.** | | | | | | | |
| --- | --- | --- | --- | --- | --- | --- | --- |
|  | **Placebo** (*n* = 16) | | | **kMCT** (*n* = 17) | | | **Δ placebo vs Δ kMCT** |
|  | **Pre** | **Post** | ***P* value*** | **Pre** | **Post** | ***P* value*** | ***P* value**† |
| PCC_L | 0.08 ± 0.03 | 0.09 ± 0.03 | 0.083 | 0.08 ± 0.03 | 0.09 ± 0.03 | 0.353 | 0.845 |
| PCC_R | 0.08 ± 0.03 | 0.09 ± 0.03 | 0.495 | 0.08 ± 0.02 | 0.09 ± 0.02 | **0.013** | 0.363 |
| PHC_L | 0.10 ± 0.03 | 0.12 ± 0.04 | 0.074 | 0.10 ± 0.04 | 0.12 ± 0.05 | **0.002** | 0.763 |
| PHC_R | 0.12 ± 0.03 | 0.14 ± 0.05 | 0.065 | 0.12 ± 0.04 | 0.15 ± 0.04 | **<0.001** | 0.510 |
| FX_L | 0.54 ± 0.07 | 0.56 ± 0.08 | 0.298 | 0.55 ± 0.06 | 0.58 ± 0.07 | **0.039** | 0.657 |
| FX_R | 0.57 ± 0.05 | 0.59 ± 0.07 | 0.375 | 0.58 ± 0.07 | 0.59 ± 0.06 | 0.430 | >0.999 |
| AF_L | 0.10 ± 0.04 | 0.12 ± 0.04 | **0.009** | 0.10 ± 0.05 | 0.13 ± 0.06 | **<0.001** | 0.533 |
| AF_R | 0.11 ± 0.04 | 0.13 ± 0.05 | **0.006** | 0.12 ± 0.07 | 0.13 ± 0.06 | **0.009** | 0.957 |
| CC2 | 0.20 ± 0.04 | 0.21 ± 0.05 | 0.073 | 0.19 ± 0.04 | 0.21 ± 0.05 | **0.020** | 0.817 |
| CC7 | 0.18 ± 0.04 | 0.20 ± 0.06 | 0.348 | 0.18 ± 0.06 | 0.20 ± 0.06 | **0.004** | 0.118 |
| IFOF_L | 0.16 ± 0.04 | 0.18 ± 0.05 | **0.018** | 0.15 ± 0.04 | 0.17 ± 0.05 | **0.001** | 0.762 |
| IFOF_R | 0.17 ± 0.04 | 0.18 ± 0.05 | **0.025** | 0.16 ± 0.05 | 0.18 ± 0.05 | **0.003** | 0.735 |
| ILF_L | 0.16 ± 0.05 | 0.18 ± 0.06 | **0.018** | 0.15 ± 0.05 | 0.17 ± 0.05 | **0.001** | 0.382 |
| ILF_R | 0.16 ± 0.04 | 0.18 ± 0.05 | **0.004** | 0.16 ± 0.05 | 0.18 ± 0.06 | **<0.001** | 0.709 |
| UF_L | 0.15 ± 0.04 | 0.17 ± 0.05 | **0.029** | 0.14 ± 0.03 | 0.16 ± 0.04 | **0.002** | 0.736 |
| UF_R | 0.16 ± 0.04 | 0.17 ± 0.05 | 0.074 | 0.15 ± 0.04 | 0.17 ± 0.04 | **0.013** | 0.606 |

Abbreviations: kMCT, ketogenic medium chain triglyceride; PCC, posterior cingulate segment of the cingulum; PHC, parahippocampal segment of the cingulum; FX, fornix; AF, arcuate fasciculus; CC2, genu of the corpus callosum; CC7, splenium of the corpus callosum; IFOF, inferior fronto-occipital fasciculus; ILF, inferior longitudinal fasciculus; UF, uncinate fasciculus.

* Statistical analysis was made using Wilcoxon signed rank test for matched pairs.

† Statistical analysis was made using Mann-Whitney test.

| **Table A.6. White matter fascicle-based glucose (FDG)* uptake before (Pre) and after (Post) the 6-month intervention.** | | | | | | | |
| --- | --- | --- | --- | --- | --- | --- | --- |
|  | **Placebo** (*n* = 16) | | | **kMCT** (*n* = 17) | | | **Δ placebo vs Δ kMCT** |
|  | **Pre** | **Post** | ***P* value**† | **Pre** | **Post** | ***P* value**† | ***P* value**‡ |
| PCC_L | 22.69 ± 3.06 | 20.78 ± 2.58 | **0.032** | 22.21 ± 2.43 | 19.67 ± 3.55 | **0.009** | 0.711 |
| PCC_R | 22.47 ± 4.71 | 20.52 ± 2.74 | 0.058 | 22.33 ± 3.63 | 19.88 ± 3.56 | **0.044** | 0.769 |
| PHC_L | 19.42 ± 3.98 | 18.85 ± 2.59 | 0.217 | 18.16 ± 3.00 | 18.11 ± 2.99 | >0.999 | 0.336 |
| PHC_R | 18.46 ± 3.40 | 19.24 ± 2.60 | >0.999 | 18.60 ± 3.08 | 17.92 ± 3.22 | 0.487 | 0.421 |
| FX_L | 14.80 ± 3.98 | 14.92 ± 2.27 | >0.999 | 12.96 ± 2.88 | 13.73 ± 4.09 | 0.517 | 0.891 |
| FX_R | 13.85 ± 3.38 | 14.13 ± 2.68 | 0.903 | 12.03 ± 3.54 | 13.81 ± 4.26 | 0.109 | 0.214 |
| AF_L | 15.37 ± 2.59 | 15.68 ± 1.51 | 0.626 | 15.32 ± 2.32 | 15.55 ± 2.35 | 0.611 | 0.399 |
| AF_R | 15.11 ± 1.70 | 16.06 ± 1.99 | 0.241 | 14.88 ± 2.50 | 15.19 ± 2.49 | 0.517 | 0.922 |
| CC2 | 15.43 ± 3.21 | 16.23 ± 1.64 | 0.903 | 15.06 ± 2.33 | 15.51 ± 2.18 | 0.431 | 0.799 |
| CC7 | 16.76 ± 2.76 | 16.38 ± 1.62 | 0.241 | 16.66 ± 2.32 | 15.89 ± 2.58 | 0.225 | 0.830 |
| IFOF_L | 16.20 ± 2.30 | 16.65 ± 1.70 | 0.626 | 16.27 ± 2.01 | 16.17 ± 2.59 | 0.782 | 0.984 |
| IFOF_R | 16.60 ± 2.20 | 17.11 ± 1.88 | 0.626 | 16.28 ± 2.74 | 16.26 ± 2.80 | 0.963 | 0.860 |
| ILF_L | 15.29 ± 2.12 | 15.78 ± 1.63 | 0.670 | 14.93 ± 1.95 | 15.10 ± 2.43 | 0.712 | 0.891 |
| ILF_R | 16.11 ± 1.99 | 16.39 ± 1.83 | 0.217 | 15.58 ± 2.57 | 15.79 ± 2.76 | 0.644 | 0.468 |
| UF_L | 16.90 ± 2.50 | 17.69 ± 1.60 | 0.715 | 17.01 ± 2.03 | 16.98 ± 2.55 | 0.854 | 0.653 |
| UF_R | 17.05 ± 2.45 | 17.80 ± 2.19 | 0.761 | 16.94 ± 2.79 | 16.96 ± 2.88 | 0.890 | 0.739 |

Abbreviations: kMCT, ketogenic medium chain triglyceride; FDG, ^18^F-fluorodeoxyglucose; PCC, posterior cingulate segment of the cingulum; PHC, parahippocampal segment of the cingulum; FX, fornix; AF, arcuate fasciculus; CC2, genu of the corpus callosum; CC7, splenium of the corpus callosum; IFOF, inferior fronto-occipital fasciculus; ILF, inferior longitudinal fasciculus; UF, uncinate fasciculus.

* Fascicle FDG metabolic rate (µmol/100 g/min).

† Statistical analysis was made using Wilcoxon matched-pairs signed rank test.

‡ Statistical analysis was made using Mann-Whitney test.
